# Enhancing the accuracy of a multivariable prediction model to identify medical patients suitable for Same Day Emergency Care services

**DOI:** 10.1101/2024.10.25.24316135

**Authors:** C. E. Atkin, S. Gallier, J. Hodson, L. Li, F. Evison, V. Reddy-Kolanu, E. Sapey

**Affiliations:** Acute Care, College of Medicine and Health, University of Birmingham, B15 2TT. Acute Medicine, University Hospitals Birmingham NHS Foundation Trust, Birmingham, B15 2GW; Head of Research Informatics, University Hospitals Birmingham NHS Foundation Trust, Birmingham, B15 2GW; Research Development & Innovation Department, University Hospitals Birmingham NHS Foundation Trust, Mindlesohn Way, Edgbaston, Birmingham, B15 2WB, UK; Department of Acute Medicine, University Hospitals Birmingham NHS Foundation Trust, Mindlesohn Way, Edgbaston, Birmingham, B15 2WB, UK; Director of PIONEER: Health Data Research UK (HDRUK) Health Data Research Hub for Acute Care, University Hospitals Birmingham NHS Foundation Trust, Edgbaston, Birmingham, B15 2GW, UK

**Author notes:** Joint first author (C. E. Atkin, S. Gallier). A. NIHR Applied Research Collaboration West Midlands, University of Birmingham, B15 2TT, UK. B. NIHR Midlands Patient Safety Research Collaboration, University of Birmingham, B15 2TT. C. NIHR Birmingham Biomedical Research Centre, University Hospitals Birmingham NHS Foundation Trust, Edgbaston, Birmingham, B15 2GW.

**Keywords:** same day emergency care, admission pathways, acute medicine

## Abstract

**Objectives:** To test the performance of the Glasgow Admission Prediction Score (GAPS) and Ambulatory Score (Amb score), and derive and validate a novel score for the identification of Emergency Department (ED) attendances suitable for treatment by Same Day Emergency Care (SDEC) services.

**Design:** Retrospective diagnostic study using routinely collected data from electronic healthcare records.

**Setting:** Three hospitals in the diverse urban setting of Birmingham, UK, between April 2023-March 2024.

**Participants:** Adult patients with an unplanned hospital attendance requiring internal medicine assessment.

**Main Outcome Measures:** Suitability for treatment by SDEC services, defined as being discharged alive with a length of stay of <12 hours (“LOS<12”).

**Results:** Data were included for 152,877 attendances, with a median age of 58 years (interquartile range: 38 to 76), and of which 54.3% were by female patients and 68.4% of White ethnicity; the outcome of LOS<12 was achieved in 45.0% (N=68,752). The GAPS and Amb score had moderate predictive accuracy, with areas under the receiver operating characteristic curve (AUROCs) of 0.741 (95% CI: 0.738 to 0.744) and 0.733 (95% CI: 0.730 to 0.736), respectively. A novel score was produced, comprising the factors from the GAPS and Amb score, as well as the National Early Warning Score 2 (NEWS2) and primary presenting complaint. When applied to an internal validation set (N=27,078), the resulting SDEC Triage Tool (SDEC-T) achieved an AUROC of 0.850 (95% CI: 0.845 to 0.854), with performance being similar across the three hospitals (AUROC range: 0.845 to 0.858).

**Conclusions:** The novel score derived within this diverse cohort has superior accuracy to the existing Amb score and GAPS for the identification of patients suitable for treatment in SDEC.

## Introduction

Nationally and globally there is increasing demand on urgent and emergency healthcare services, with rising presentations to hospital Emergency Departments (EDs).(1) This has led to overcrowding in acute care services, long delays and poor outcomes for patients.(2) Most patients requiring onwards referral and assessment have a medical issue requiring review by general and acute internal medical teams.(3) Annual benchmarking audits by the Society for Acute Medicine suggest performance against key quality indicators is deteriorating; namely measuring patient acuity on admission, assessment by a clinically-competent clinical decision maker within four hours of arrival, and assessment by a consultant physician within six hours (daytime) or 14 hours (out of hours) of admission to hospital.(4,5) To try to address this, healthcare organisations have developed new pathways which aim to streamline access to acute medical teams, to expedite clinical decision making with an aim of reducing delays and improving patient experience. One such development is Same Day Emergency Care (SDEC). SDEC services provide patients that would otherwise have required hospital admission with specialist assessment and management without admission to an inpatient bed.(6) Services are ideally located close to acute medical units (AMUs), and deliver assessment within trolley or chair spaces, rather than beds. Under the UK’s National Health Service (NHS) Long Term Plan, released in 2019, NHS England mandated that every acute hospital with a type 1 ED (consultant-led 24 hour service) will provide SDEC services.(7,8) There is an expectation that a third of all medical attendances can be managed without overnight admission, and that this could be increased from a fifth in 2018 through the increased provision of SDEC.(7) As a proportion of patients assessed within SDEC will be found to require admission, this necessitates SDEC being utilised for more than 40% of unplanned medical attendances.(6) Key to meeting these ambitious targets is the correct identification of patients likely to benefit from these new pathways, as early as possible during the admission process. Bringing patients into SDEC who then require admission interrupts patient flow and can place patients at risk of harm if their care needs exceed those which can be provided in a seated SDEC area. Conversely, bringing patients into SDEC who could be discharged more rapidly from EDs creates delays in care without discernible benefit.(9) Presently, two risk stratification tools have been deployed to identify patients suitable for treatment by SDEC services in the UK: (10) the Glasgow Admission Prediction Score (GAPS) and the Ambulatory Score (Amb score).(11) These scoring systems are both designed for use early within the assessment pathway, combining multiple patient features to predict the likelihood of same-day discharge (Amb score) or admission to hospital (GAPS). The GAPS was originally derived and validated in Glasgow,(12) with further validation in Sheffield;(13) local evaluation to determine valid cut-offs was advised by the authors. The Amb score was designed to identify medical patients likely to be discharged within 12 hours; the original validation took place at a single centre in Wales,(14) and independent evaluation has since been performed in Taunton and Malta.(15,16)

Despite the Amb score being supported by the Royal College of Physicians,(17) its uptake by hospitals across the UK has been variable, and reviews of national AMU and SDEC provision describe considerable variability in size, patient flows, and selection tools used to identify patients and staffing levels.(4,18) A potential reason for the variable use of Amb score and GAPS nationally is their inconsistent performance when used in different settings(10). A recent study including 7,365 patient attendances from a diverse urban population found that implementing the Amb score or GAPS to select patients potentially suitable for review in SDEC would have resulted in >45% of patients assessed in SDEC subsequently requiring an inpatient admission.(19)

Developing tools that can accurately identify patients suitable for SDEC in different settings would be of great benefit. Ensuring these tools can be deployed as soon as possible after presentation to an ED is critical, as those which require information based on imaging or blood results may not be available for several hours after presentation, delaying access to the most beneficial care pathway. Building tools which feel helpful and relevant to clinical staff is also key, to help with their adoption into clinical care.

## Methods

### Aims and objectives

The aims of the study were fivefold:

1. To understand what healthcare professionals involved in selecting patients for SDEC services or receiving/reviewing patients for SDEC, policy makers and members of the public who may require these services, required from a selection tool.
2. To assess the performance of the Amb score and GAPS for the identification of patients likely to be suitable for SDEC in an urban and diverse population.
3. To develop a novel tool to identify patients likely to be suitable for SDEC using routinely collected data available on initial presentation to hospital.
4. To validate this novel tool, and compare performance to the Amb score and GAPS.
5. To gain feedback on the novel tool and how it might be used by healthcare professionals involved in selecting patients for SDEC services or receiving/reviewing patients for SDEC.

### Ethics

The study was approved by the East Derby Research Ethics Committee (reference 20/EM/0158) and Health Research Authority (reference number 279353). This retrospective study was conducted in accordance with the Transparent Reporting of a Multivariable Prediction Model for Individual Prognosis or Diagnosis (TRIPOD) guidelines.(20)

### Public and Patient Involvement

Patients and the public were involved from the outset of this study, with initial discussions and presentations held with patient groups utilising acute care services prior to obtaining ethical approval. These sessions enabled co-development of the research questions and outcome measures, shaping and refining the study’s design. Patients and public representatives participated in workshops where current selection processes for SDEC were discussed and where the proposed novel tool was introduced.

### Workshop structure and theme capture

Two virtual workshops were organised, with attendance open to healthcare professionals working in Emergency Medicine, Primary Care and Acute Internal Medicine (with no professional background restrictions); policy makers involved in designing SDEC services; and to patients and members of the public. In the first scoping workshop, healthcare professionals were invited to describe the tools they currently use for selection of patients for SDEC, followed by a structured discussion based around four headings: what tools were currently used; what were their strengths and limitations; what would an optimal tool include; and how would it be used. These were discussed from the perspective of healthcare professionals, policy makers and then patients/members of the public. The discussion was recorded and transcribed, and analysed for themes, which were considered when producing the novel tool. A second evaluation workshop was then organised with the same members, to present the novel tool, and discuss its potential utility across different care settings, including onward evaluation.

### Setting

The data study included three Birmingham hospitals within the University Hospitals Birmingham NHS Foundation Trust, which provide urgent and emergency care to a diverse population of 2.2 million people in the West Midlands area, namely Birmingham Heartlands Hospital (BHH), the Queen Elizabeth Hospital Birmingham (QEHB), and Good Hope Hospital (GHH).

### Study population

All attendances to one of the three EDs by adults (aged 16+ years) between 1^st^ April 2023 and 31^st^ March 2024 (inclusive) were retrospectively identified. Patients who were referred directly to one of AMUs without first attending the ED were also identified and included in the study. These patients can be referred from several sources, namely the NHS 111 Service, General Practitioners (GPs), or via the ambulance service.

Attendances that did not necessitate assessment by the internal medical team were then excluded, on the basis that these would not be considered for SDEC, specifically patients who:

- Were referred from the ED to a surgical specialty, psychiatry, stroke medicine, trauma, or the Intensive Care Unit.
- Presented with suspected venous thromboembolism (VTE) and attended the VTE outpatient clinic within 14 days of the ED attendance, as the optimum management for these patients is via a well-established community pathway.
- Attended AMU to receive a regular, planned administration of intravenous (IV) antibiotics.
- Were only referred to a fracture clinic, a knee or shoulder clinic, or for physiotherapy.
- Were discharged home from the ED without an onwards referral.

Further details of the exclusion criteria are summarised in ***Supplementary Figure 1***.

Validation of the existing GAPS and Amb score tools were initially performed for the overall population. Prior to the derivation of the novel tool, the cohort was then divided into two subsets. Specifically, 80% of admissions were randomly selected to make up the derivation set, from which the tool was produced, with the remaining 20% comprising a validation set, which was used for internal validation of the tool.

### Data extraction

Data were extracted from the electronic health records (EHR) system. Patient demographics comprised age, sex and self-reported ethnicity, with the source of the ED referral (e.g. self-referral or referred by GP) and the mode of arrival (e.g. self-presentation or ambulance) also recorded. Any previous inpatient stays at one of the three study hospitals within either 30 days or one year of the ED attendance were identified. The primary presenting complaint recorded at triage was extracted and grouped into broader categories for analysis; this was not recorded in patients directly attending the AMU. The triage category (standard-, urgent- or immediate-level care) was additionally recorded and mapped to the Manchester Triage Score to calculate the GAPS; patients directly attending the AMU were treated as being triaged to standard-level care.(21)

The first sets of observations recorded within six hours of arrival were then extracted, to use in the calculation of the Modified Early Warning Score (MEWS), National Early Warning Score (NEWS) and NEWS2 scores. All prescriptions within six hours of ED attendance were reviewed, to identify any that were administered via an IV route. The dates and times of arrival at the ED and discharge from hospital (to the nearest minute) were then extracted and used to calculate the length of stay (LOS).

### Primary outcome measure

Since the suitability for treatment in SDEC could not be readily quantified using retrospectively collected data, the LOS was used as a surrogate outcome. Attendances were deemed suitable for SDEC if patients were discharged alive with a length of stay of <12 hours (referred to subsequently as “LOS<12”), on the basis that they could potentially have been treated and discharged on the same day. Attendances with a LOS of ≥12 hours, or where patients died within 12 hours of arrival did not achieve the outcome of LOS<12, and so were deemed to be unsuitable for SDEC. This is consistent with the outcome used to signify SDEC suitability in the derivation of the Amb score.(14)

### Calculation of existing predictive scores

To calculate the GAPS and Amb scores, it was necessary to make some assumptions due to the limitations of the retrospective data. For the Amb score, it was not possible to identify whether patients had *“access to personal transport/can take public transport”*. However, the three study hospitals are well served by public transport and provide transport to patients, as required. As such, it was assumed that all attendances fulfilled this criterion, and so all cases were assigned two points. For the component regarding the anticipation of IV treatment, it was assumed that such treatments administered within six hours of arrival would likely have been anticipated; hence, these cases were assigned zero points, with two points assigned in the remainder. Finally, for the components of both scores relating to previous inpatient admissions, since data were collected from local EHR systems, only inpatient stays at the study hospitals could be identified; hence, any admissions to other hospitals could not be identified and so were not considered when calculating the scores. The resulting lookup tables used to calculate the two scores are reported in ***Table 1***.

**Table 1.** Calculation of GAPS and Amb scores.

| GAPS |  | Amb Score |  |
| --- | --- | --- | --- |
| Component | Points | Component | Points |
| Age | 1 per Decade | Age |  |
| Inpatient in Previous Year <sup>a</sup> |  | <80 Years | 0 |
| No | 0 | ≥80 Years | -0.5 |
| Yes | 5 | Sex |  |
| Referred by GP |  | Female | 0 |
| No | 0 | Male | -0.5 |
| Yes | 10 | Inpatient in Previous 30 Days <sup>a</sup> |  |
| Arrived in Ambulance |  | No | 1 |
| No | 0 | Yes | 0 |
| Yes | 5 | Acutely Confused |  |
| NEWS | 1 per Point | No | 2 |
| Triage Category |  | Yes | 0 |
| Standard | 5 | MEWS |  |
| Urgent | 10 | 0 | 1 |
| Immediate | 20 | >0 | 0 |
|  |  | IV Treatment Within 6 Hours <sup>b</sup> |  |
|  |  | No | 2 |
|  |  | Yes | 0 |
|  |  | Access to Transportation <sup>c</sup> |  |
|  |  | No | 0 |
|  |  | Yes | 2 |

**Legend.** <sup>a</sup> Only previous admissions to one of the three study hospitals were included. <sup>b</sup> In the original Amb score, this component related to whether IV treatment was “anticipated”; due to the limitations of the retrospective study design, the administration of IV treatment within six hours of attendance was used as a surrogate marker. <sup>c</sup> All patients were assumed to have access to transportation, and so were assigned two points. Amb Score: Ambulatory Score, GAPS: Glasgow Admission Prediction Score, GP: General Practitioner, IV: Intravenous, MEWS: Modified Early Warning Score, NEWS: National Early Warning Score

### Derivation of a novel tool

A novel tool was then produced, with all of the component factors from the Amb score and GAPS considered for inclusion, with the exception of access to transportation. The MEWS and NEWS, which were used in the Amb score and GAPS, respectively, were replaced with NEWS2, as this is now the acuity score mandated for use in NHS hospitals. (22) The novel score additionally considered the primary presenting complaint for inclusion.

### Statistical methods

Since the observations used to calculate the NEWS/MEWS and, hence, the GAPS and Amb score were not available for all patients, comparisons were initially made between those attendances for which these acuity scores were and were not calculable, to identify any evidence of selection bias. Mann-Whitney U tests were used for continuous or ordinal factors, with nominal factors analysed using Chi-square tests. The predictive accuracy of the GAPS and Amb scores, with respect to LOS<12, was then assessed using receiver operating characteristic (ROC) curves and quantified using the area under the ROC curve (AUROC).

For the derivation set, separate univariable binary logistic regression models were produced to assess the associations between each factor and LOS<12. For continuous factors, the goodness-of-fit of the resulting models were assessed graphically. A multivariable analysis was then performed, which used backwards-stepwise variable selection (removal at p>0.1) to produce a parsimonious model comprising factors that were independently predictive of LOS<12. The resulting model was then converted into a predictive score. For nominal variables, the reference categories for factors were changed, such that effects were in a positive direction, where possible. The natural logs of the odds ratios (LnORs) of each factor were then multiplied by a constant, to give values of a reasonable magnitude and to minimise the impact of rounding. The resulting values were then rounded to the nearest integer, to assign a point score to each component.

The optimal threshold for prediction of LOS<12 then identified for the novel tool, as well as for the GAPS and Amb score, based on the value that maximised the Youden’s J statistic. The novel tool was then applied to the validation set, with the predictive accuracy quantified by the AUROC as well as measures of classification accuracy (sensitivity, specificity and positive/negative predictive values), based on the previously defined threshold. The validation analysis of the novel tool was also repeated for each of the three hospitals separately, with the GAPS and Amb scores also assessed on the validation set, for reference.

All analyses were performed using IBM SPSS v29 (IBM Corp. Armonk, NY), with p<0.05 deemed to be indicative of statistical significance throughout. Clustering by hospital was not considered when deriving the novel tool; however, validation was additionally performed for each hospital separately, to identify any variability in the performance of the novel tool.

Cases with missing data were excluded from univariable analyses of the affected factors, with multivariable analyses using a complete-cases approach. Continuous variables were not found to be normally distributed, and so are summarised using medians and interquartile ranges (IQRs).

## Results

### Scoping Workshop

The initial workshop was attended by 31 people in total, from England, Scotland and Wales, with clinical and non-clinical professional backgrounds including doctors; nurses and Advanced Clinical Practitioners; and attendees from NHSE, acute medicine, emergency medicine and surgery; as well as members of the public. There was consensus that a formal tool to help identify patients suitable for SDEC would be helpful to improve care and standardise practice. The majority of centres represented in the workshop did not have any set criteria for selecting patients for SDEC. Those that did used the NEWS2 score, the Clinical Frailty Scale and one centre used the Amb score. In discussions of what information may be helpful in selecting patients for SDEC, it was agreed that only data which was readily available on admission should be included. Imaging and blood results (even those potentially available via point of care testing) should be excluded, due to difficulty in interpretation by triaging staff. Most centres described how certain presenting complaints were used to help triage to SDEC, and these factors could be included. The most common themes around deployment of any tool concerned ease of use. Specifically, that the tool would comprise factors that were simple to measure, so that it could be utilised as early as possible during the presentation, including during conveyance from the community (i.e. bypassing the ED). In addition, it was suggested that the tool itself should be simple to calculate, to reduce the risk of calculation errors, and simplify potential implementation into electronic systems. It was recognised that the level of digital maturity varied considerably within the NHS, which impacted the ability to deploy tools which required technical infrastructure, including large language models. These factors were considered in tool development.

### Cohort characteristics

Data were available for N=152,877 attendances, with patients having a median age of 58 (IQR: 38 to 76) years and the most common primary presenting complaints being chest pain (18.0%) and shortness of breath (14.1%). The median length of stay was 17 (IQR: 5 to 101) hours, with 45.0% (N=68,752) of attendances achieving the outcome of LOS<12; further details of the cohort are summarised in ***Table 2***. Comparisons between the three study hospitals found large differences in patient demographics, with GHH having a preponderance of attendances for older patients (median age: 67 years) of White ethnicity (86.6%), whilst patients attending BHH were considerably younger (median: 54 years) and more ethnically diverse (58.2% White).

**Table 2.**
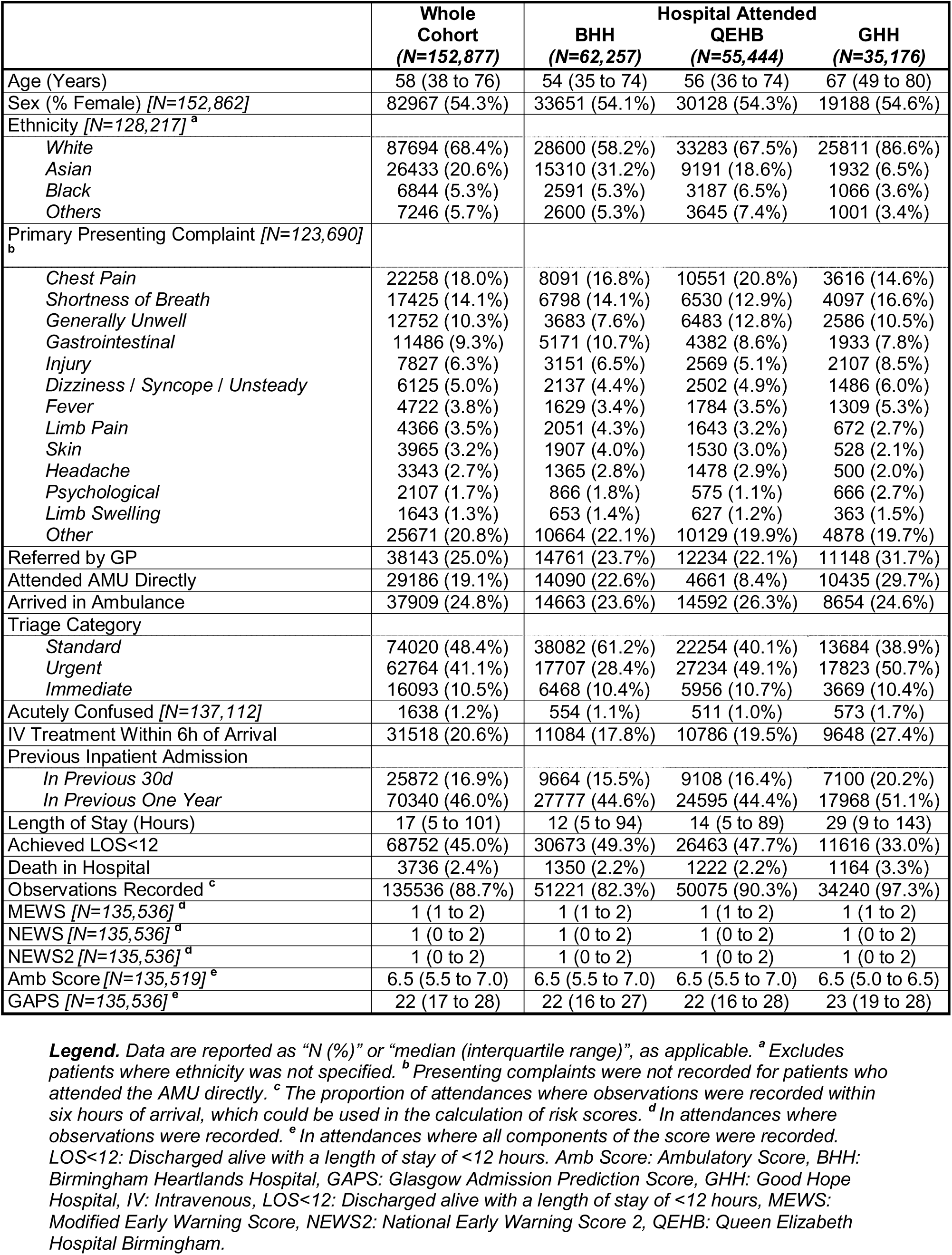
Characteristics of primary cohort.

|  | Whole Cohort<br>(N=152,877) | Hospital Attended |  |  |
| --- | --- | --- | --- | --- |
|  |  | BHH<br>(N=62,257) | QEHB<br>(N=55,444) | GHH<br>(N=35,176) |
| Age (Years) | 58 (38 to 76) | 54 (35 to 74) | 56 (36 to 74) | 67 (49 to 80) |
| Sex (% Female) [N=152,862] | 82967 (54.3%) | 33651 (54.1%) | 30128 (54.3%) | 19188 (54.6%) |
| Ethnicity [N=128,217]<sup>a</sup> |  |  |  |  |
| White | 87694 (68.4%) | 28600 (58.2%) | 33283 (67.5%) | 25811 (86.6%) |
| Asian | 26433 (20.6%) | 15310 (31.2%) | 9191 (18.6%) | 1932 (6.5%) |
| Black | 6844 (5.3%) | 2591 (5.3%) | 3187 (6.5%) | 1066 (3.6%) |
| Others | 7246 (5.7%) | 2600 (5.3%) | 3645 (7.4%) | 1001 (3.4%) |
| Primary Presenting Complaint [N=123,690]<sup>b</sup> |  |  |  |  |
| Chest Pain | 22258 (18.0%) | 8091 (16.8%) | 10551 (20.8%) | 3616 (14.6%) |
| Shortness of Breath | 17425 (14.1%) | 6798 (14.1%) | 6530 (12.9%) | 4097 (16.6%) |
| Generally Unwell | 12752 (10.3%) | 3683 (7.6%) | 6483 (12.8%) | 2586 (10.5%) |
| Gastrointestinal | 11486 (9.3%) | 5171 (10.7%) | 4382 (8.6%) | 1933 (7.8%) |
| Injury | 7827 (6.3%) | 3151 (6.5%) | 2569 (5.1%) | 2107 (8.5%) |
| Dizziness / Syncope / Unsteady | 6125 (5.0%) | 2137 (4.4%) | 2502 (4.9%) | 1486 (6.0%) |
| Fever | 4722 (3.8%) | 1629 (3.4%) | 1784 (3.5%) | 1309 (5.3%) |
| Limb Pain | 4366 (3.5%) | 2051 (4.3%) | 1643 (3.2%) | 672 (2.7%) |
| Skin | 3965 (3.2%) | 1907 (4.0%) | 1530 (3.0%) | 528 (2.1%) |
| Headache | 3343 (2.7%) | 1365 (2.8%) | 1478 (2.9%) | 500 (2.0%) |
| Psychological | 2107 (1.7%) | 866 (1.8%) | 575 (1.1%) | 666 (2.7%) |
| Limb Swelling | 1643 (1.3%) | 653 (1.4%) | 627 (1.2%) | 363 (1.5%) |
| Other | 25671 (20.8%) | 10664 (22.1%) | 10129 (19.9%) | 4878 (19.7%) |
| Referred by GP | 38143 (25.0%) | 14761 (23.7%) | 12234 (22.1%) | 11148 (31.7%) |
| Attended AMU Directly | 29186 (19.1%) | 14090 (22.6%) | 4661 (8.4%) | 10435 (29.7%) |
| Arrived in Ambulance | 37909 (24.8%) | 14663 (23.6%) | 14592 (26.3%) | 8654 (24.6%) |
| Triage Category |  |  |  |  |
| Standard | 74020 (48.4%) | 38082 (61.2%) | 22254 (40.1%) | 13684 (38.9%) |
| Urgent | 62764 (41.1%) | 17707 (28.4%) | 27234 (49.1%) | 17823 (50.7%) |
| Immediate | 16093 (10.5%) | 6468 (10.4%) | 5956 (10.7%) | 3669 (10.4%) |
| Acutely Confused [N=137,112] | 1638 (1.2%) | 554 (1.1%) | 511 (1.0%) | 573 (1.7%) |
| IV Treatment Within 6h of Arrival | 31518 (20.6%) | 11084 (17.8%) | 10786 (19.5%) | 9648 (27.4%) |
| Previous Inpatient Admission |  |  |  |  |
| In Previous 30d | 25872 (16.9%) | 9664 (15.5%) | 9108 (16.4%) | 7100 (20.2%) |
| In Previous One Year | 70340 (46.0%) | 27777 (44.6%) | 24595 (44.4%) | 17968 (51.1%) |
| Length of Stay (Hours) | 17 (5 to 101) | 12 (5 to 94) | 14 (5 to 89) | 29 (9 to 143) |
| Achieved LOS<12 | 68752 (45.0%) | 30673 (49.3%) | 26463 (47.7%) | 11616 (33.0%) |
| Death in Hospital | 3736 (2.4%) | 1350 (2.2%) | 1222 (2.2%) | 1164 (3.3%) |
| Observations Recorded<sup>c</sup> | 135536 (88.7%) | 51221 (82.3%) | 50075 (90.3%) | 34240 (97.3%) |
| MEWS [N=135,536]<sup>d</sup> | 1 (1 to 2) | 1 (1 to 2) | 1 (1 to 2) | 1 (1 to 2) |
| NEWS [N=135,536]<sup>d</sup> | 1 (0 to 2) | 1 (0 to 2) | 1 (0 to 2) | 1 (0 to 2) |
| NEWS2 [N=135,536]<sup>d</sup> | 1 (0 to 2) | 1 (0 to 2) | 1 (0 to 2) | 1 (0 to 2) |
| Amb Score [N=135,519]<sup>e</sup> | 6.5 (5.5 to 7.0) | 6.5 (5.5 to 7.0) | 6.5 (5.5 to 7.0) | 6.5 (5.0 to 6.5) |
| GAPS [N=135,536]<sup>e</sup> | 22 (17 to 28) | 22 (16 to 27) | 22 (16 to 28) | 23 (19 to 28) |

**Legend.** Data are reported as “N (%)” or “median (interquartile range)”, as applicable. <sup>a</sup> Excludes patients where ethnicity was not specified. <sup>b</sup> Presenting complaints were not recorded for patients who attended the AMU directly. <sup>c</sup> The proportion of attendances where observations were recorded within six hours of arrival, which could be used in the calculation of risk scores. <sup>d</sup> In attendances where observations were recorded. <sup>e</sup> In attendances where all components of the score were recorded. LOS<12: Discharged alive with a length of stay of <12 hours. Amb Score: Ambulatory Score, BHH: Birmingham Heartlands Hospital, GAPS: Glasgow Admission Prediction Score, GHH: Good Hope Hospital, IV: Intravenous, LOS<12: Discharged alive with a length of stay of <12 hours, MEWS:

### Predictive accuracy of GAPS and Amb scores

The GAPS and Amb score incorporate the MEWS and NEWS score, respectively. However, these were unavailable for 11.3% (N=17,341) of the cohort, due to no observations being recorded within six hours of arrival; hence, these attendances were excluded from analysis of the two scores . Comparisons of these excluded attendances to the remainder of the cohort indicated selection bias, with patients being younger, less likely to be triaged as requiring urgent or immediate care, and having a considerably shorter length of stay, with a median of less than one hour, usually being discharged directly from ED (***Supplementary Table 1***). The Amb score was additionally incalculable for N=12 attendances where patient’s sex was not recorded, and N=5 with no data relating to acute confusion. After excluding these attendances, the GAPS was calculated for the remaining N=135,536 attendances, with a median score of 22 (IQR: 17 to 28). The Amb score was calculated for N=135,519 attendances had a median score of 6.5 (IQR: 5.5 to 7.0)

Both scores were found to be significantly predictive of LOS<12 (both p<0.001), with the GAPS having marginally superior performance, yielding an AUROC of 0.741 (95% CI: 0.738 to 0.744), compared to 0.733 (95% CI: 0.730 to 0.736) for the Amb score (***Figure 1***).

**Figure 1.**
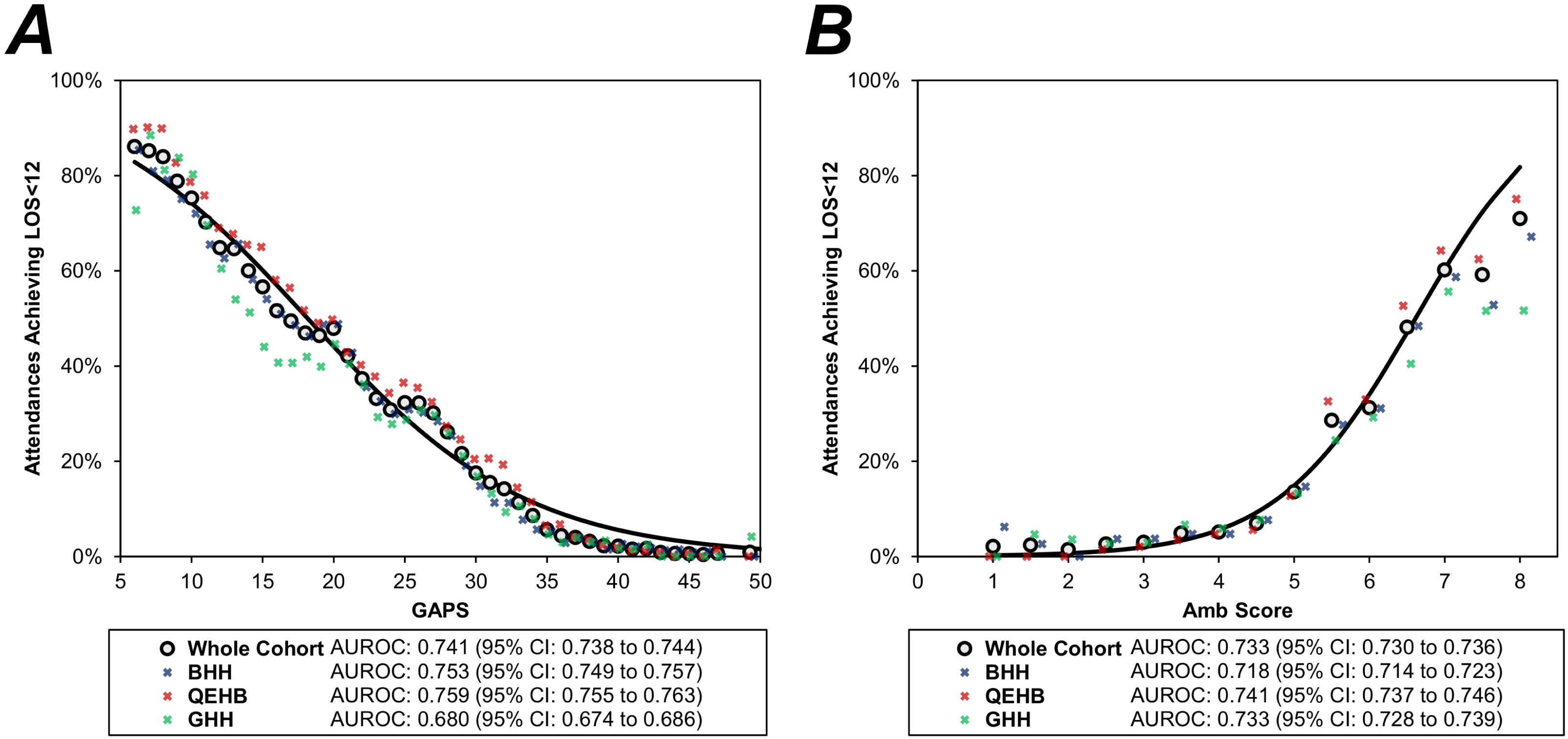
Associations between the GAPS/Amb score and LOS<12. **Legend**. Points represent the rate of LOS<12 for each value of the score and are plotted for the whole cohort of attendances for which the score were calculable (N=135,536 for the GAPS and N=135,519 for the Amb score), as well as separately for each of the three hospitals. Points are plotted with jitter on the x-axis, to prevent excessive overlaps; for the GAPS, the final point combines scores of 48-54, due to small sample sizes, and is plotted at the mean score within this interval. Trend lines are from binary logistic regression models on the admission-level data for the whole cohort, with the risk score as a continuous covariate. Amb Score: Ambulatory Score, AUROC: Area under the receiver operating characteristic curve, BHH: Birmingham Heartlands Hospital, GAPS: Glasgow Admission Prediction Score, GHH: Good Hope Hospital, LOS<12: LOS<12: Discharged alive with a length of stay of <12 hours, QEHB: Queen Elizabeth Hospital Birmingham.

Subgroup analyses of the three hospitals found the GAPS to have notably poorer performance when applied to the subgroup of patients attending GHH (AUROC: 0.680 vs. 0.759 for QEHB), whilst performance of the Amb score was more consistent across hospitals, with AUROCs ranging from 0.718 to 0.741.

### Producing novel risk score: the SDEC Triage Tool (SDEC-T)

N=122,302 admissions comprised a derivation set, which was used to produce a novel risk score. When assessing whether patients had a recent inpatient admission, only those occurring in the previous year (rather than 30 days) were considered, as this had superior predictive accuracy for LOS<12 (AUROC: 0.625 vs. 0.566). In addition, to minimise exclusions due to missing data, the subgroup of attendances where the primary presenting complaint was not recorded due to attending the AMU directly were combined with the “other” presentations. On multivariable analysis, all factors were found to be significant independent predictors of LOS<12 and selected for inclusion by the stepwise procedure, with the exception of acute confusion (***Table 3***). Whilst this had been associated with a significantly lower rate of LOS<12 on univariable analysis (6.8% vs. 39.4%, p<0.001), the low prevalence of acute confusion and correlation with the NEWS2 score meant that it did not contribute sufficiently to be included in the parsimonious model.

**Table 3.** Multivariable analyses of LOS<12.

|  | LOS<br><12 | Univariable Models<br>OR (95% CI) | Multivariable Model<br>OR (95% CI) |
| --- | --- | --- | --- |
| Age (per Decade) <sup>a</sup> | - <sup>a</sup> | 0.684 (0.680 to 0.688) | 0.794 (0.788 to 0.800) |
| Sex |  |  |  |
| Male | 42.3% | 1 | 1 |
| Female | 47.2% | 1.221 (1.193 to 1.249) | 1.195 (1.159 to 1.232) |
| Referred by GP |  |  |  |
| No | 40.5% | 1 | 1 |
| Yes | 58.6% | 2.082 (2.028 to 2.138) | 1.157 (1.109 to 1.207) |
| Arrived in Ambulance |  |  |  |
| No | 52.5% | 1 | 1 |
| Yes | 22.0% | 0.255 (0.247 to 0.263) | 0.669 (0.642 to 0.696) |
| Inpatient in Previous Year |  |  |  |
| No | 56.4% | 1 | 1 |
| Yes | 31.5% | 0.356 (0.348 to 0.364) | 0.625 (0.606 to 0.645) |
| NEWS2 (per Point) <sup>a</sup> | - <sup>a</sup> | 0.615 (0.609 to 0.621) | 0.781 (0.771 to 0.790) |
| IV Treatment Within 6h |  |  |  |
| No | 54.6% | 1 | 1 |
| Yes | 7.8% | 0.070 (0.067 to 0.074) | 0.223 (0.211 to 0.235) |
| Acutely Confused |  |  |  |
| No | 39.4% | 1 | - |
| Yes | 6.8% | 0.112 (0.091 to 0.139) | NS |
| Triage Category |  |  |  |
| Standard | 71.2% | 1 | 1 |
| Urgent | 24.5% | 0.131 (0.128 to 0.135) | 0.266 (0.256 to 0.276) |
| Immediate | 4.3% | 0.018 (0.017 to 0.020) | 0.117 (0.106 to 0.129) |
| Primary Presenting Complaint <sup>b</sup> |  |  |  |
| Fever | 10.3% | 0.090 (0.081 to 0.100) | 0.667 (0.559 to 0.796) |
| Psychological | 16.2% | 0.152 (0.133 to 0.173) | 0.433 (0.367 to 0.510) |
| Shortness of Breath | 16.3% | 0.153 (0.146 to 0.161) | 0.969 (0.903 to 1.041) |
| Generally Unwell | 29.4% | 0.327 (0.312 to 0.343) | 0.948 (0.887 to 1.012) |
| Dizziness/Syncope/Unsteady | 29.4% | 0.328 (0.308 to 0.350) | 0.850 (0.781 to 0.926) |
| Injury | 35.0% | 0.423 (0.400 to 0.447) | 1.155 (1.067 to 1.251) |
| Limb Swelling | 47.4% | 0.709 (0.635 to 0.791) | 1.658 (1.448 to 1.899) |
| Gastrointestinal | 47.9% | 0.722 (0.690 to 0.755) | 0.831 (0.775 to 0.891) |
| Chest Pain | 55.4% | 0.978 (0.944 to 1.013) | 2.050 (1.949 to 2.155) |
| Other / Direct to AMU <sup>b</sup> | 56.0% | 1 | 1 |
| Headache | 59.1% | 1.136 (1.050 to 1.229) | 1.023 (0.925 to 1.131) |
| Limb Pain | 59.5% | 1.156 (1.078 to 1.240) | 1.510 (1.370 to 1.664) |
| Skin | 72.0% | 2.020 (1.865 to 2.186) | 1.382 (1.236 to 1.546) |

The resulting model was then converted to a novel risk score, the SDEC-T, the lookup table for which is reported in ***Table 4***. Comparison of the novel score to the existing GAPS and Amb score highlighted several similarities. Like the GAPS, SDEC-T assigned one point per decade of age or per point of the NEWS/NEWS2 score, albeit negative points, since the GAPS was intended to predict extended rather than short hospital stays. SDEC-T also included whether patients were inpatients in the previous year, arrival by ambulance and the triage category, but assigned points which gave approximately half the weight to these, compared to the GAPS. The only major discrepancy was that SDEC-T identified patients referred by their GP as being more likely to have LOS <12, which was the converse of the GAPS. Of the additional factors from the Amb score, female sex and IV treatment not being anticipated were included in the novel score and assigned similar weights, although acute confusion was excluded, as previously described. Of the primary presenting complaints considered, SDEC-T found the likelihood of LOS<12 to be lowest in admissions with psychological symptoms, and highest in those presenting with either pain or swelling of the limbs, or with chest pain.

**Table 4.** SDEC-T lookup table.

| Component | Points |
| --- | --- |
| Age | *Minus 1 per Decade* |
| Sex |  |
| *Male* | 0 |
| *Female* | 1 |
| Referred by GP |  |
| *No* | 0 |
| *Yes* | 1 |
| Arrived in Ambulance |  |
| *No* | 2 |
| *Yes* | 0 |
| Inpatient in Previous Year |  |
| *No* | 2 |
| *Yes* | 0 |
| NEWS2 | *Minus 1 per Point* |
| IV Treatment Anticipated |  |
| *No* | 6 |
| *Yes* | 0 |
| Triage Category |  |
| *Standard* | 9 |
| *Urgent* | 4 |
| *Immediate* | 0 |
| Primary Presenting Complaint |  |
| *Not Listed Below* | 0 |
| *Psychological* | *Minus 3* |
| *Fever* | *Minus 2* |
| *Gastrointestinal* | *Minus 1* |
| *Dizziness/Syncope/Unsteady* | *Minus 1* |
| *Injury* | 1 |
| *Skin* | 1 |
| *Limb Pain* | 2 |
| *Limb Swelling* | 2 |
| *Chest Pain* | 3 |

### SDEC-T validation

Prior to validating SDEC-T, the optimal threshold was identified using the admissions from the derivation set. This identified a score of ≥9 to be the best predictor of LOS<12; similar analysis of the existing scores returned thresholds of ≤21 for the GAPS and ≥6.5 for the Amb score.

The scores were then applied to the N=27,078 (88.6%) admissions from the validation set for which all three scores were calculable. This found SDEC-T to be strongly predictive of LOS<12, with an AUROC of 0.850 (95% CI: 0.845 to 0.854), which was similar across the three hospitals (range: 0.845 to 0.858, ***Figure 2***). Using a threshold of ≥9 would result in 49.9% of attendances being deemed potentially suitable for SDEC (i.e. predicted to achieve the outcome of LOS<12) and yield 84.6% sensitivity and 72.4% specificity (***Table 5***). SDEC-T outperformed both the GAPS and Amb scores, which returned AUROCs of 0.737 (95% CI: 0.731 to 0.743) and 0.734 (95% CI: 0.728 to 0.740), respectively, when applied to the validation set. Subgroup analysis of the novel score by subgroups defined by the NEWS2 found the performance to be similar in those with NEWS2≤4 (N=24,338; AUROC: 0.828; 95% CI: 0.823 to 0.833) and with NEWS2>4 (N=2,740; AUROC: 0.826; 95% CI: 0.782 to 0.869).

**Figure 2.**
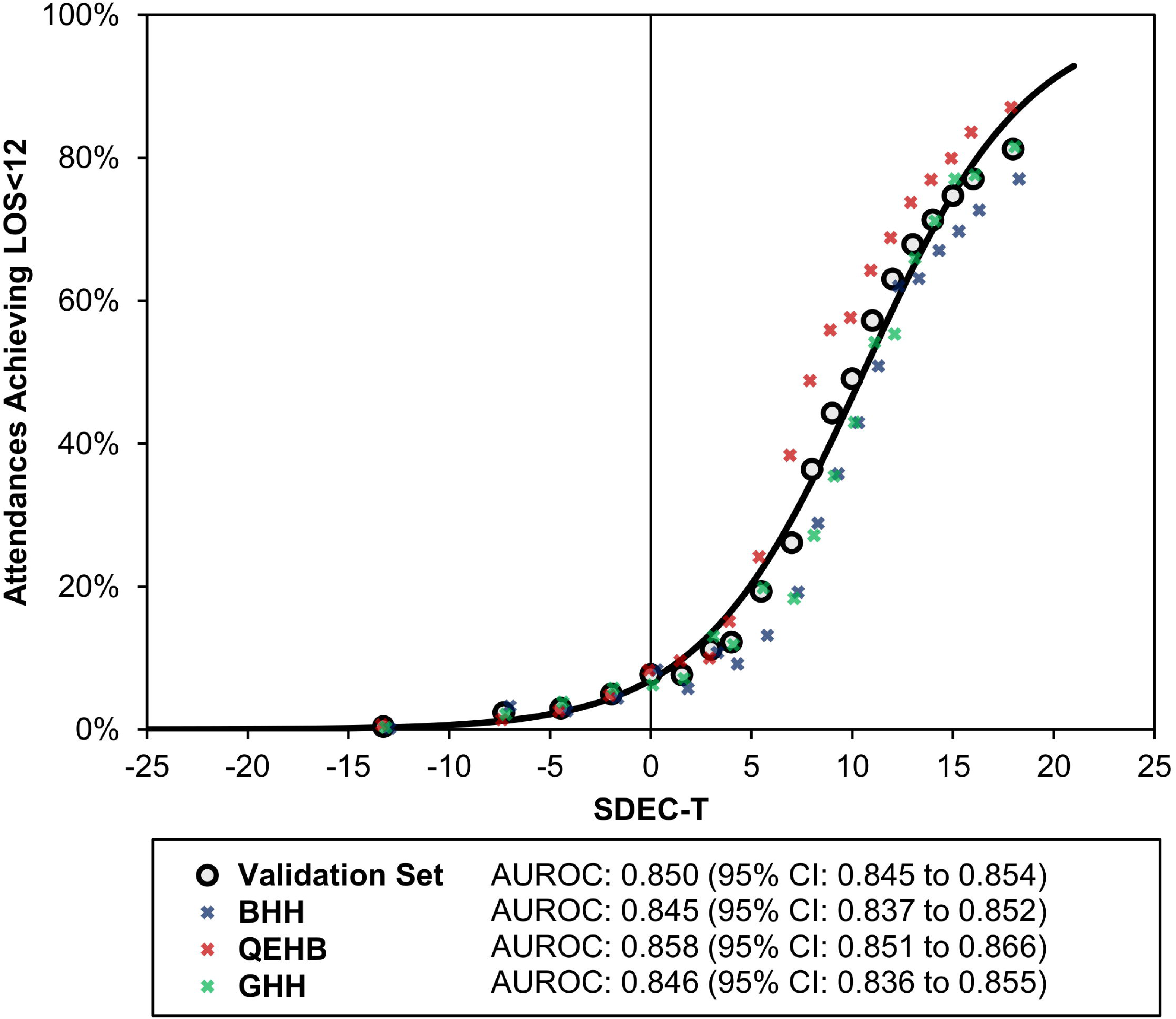
Association between SDEC-T and LOS<12 for the validation set. **Legend.** Analyses are based on attendances from the validation set for which the SDEC-T was calculable (N=27,078). Attendances were divided into 20 subgroups, based on the percentiles of the SDEC-T, with points representing the rate of LOS<12 within each subgroup, plotted at the mean score within the subgroup. Points are plotted for the validation set as a whole, as well as separately for each of the three hospitals with jitter on the x-axis, to prevent excessive overlaps. The trend line is from a binary logistic regression model on the admission-level data for the whole validation set, with the SDEC-T as a continuous covariate. AUROC: Area under the receiver operating characteristic curve, BHH: Birmingham Heartlands Hospital, GHH: Good Hope Hospital, LOS<12: Discharged alive with a length of stay of <12 hours, QEHB: Queen Elizabeth Hospital Birmingham, SDEC-T: SDEC Triage Tool.

**Table 5.** Predictive accuracy of SDEC-T and existing tools on the validation set.

| Score (Threshold) | AUROC (95% CI) | Potentially Suitable For SDEC <sup>a</sup> | Sensitivity | Specificity | Positive Predictive Value | Negative Predictive Value |
| --- | --- | --- | --- | --- | --- | --- |
| SDEC-T (≥9) | 0.850 (0.845 to 0.854) | 49.9% (13500/27078) | 84.6% (8937/10565) | 72.4% (11950/16513) | 66.2% (8937/13500) | 88.0% (11950/13578) |
| BHH | 0.845 (0.837 to 0.852) | 54.6% (5496/10066) | 89.5% (3501/3913) | 67.6% (4158/6153) | 63.7% (3501/5496) | 91.0% (4158/4570) |
| QEHB | 0.858 (0.851 to 0.866) | 50.4% (5121/10158) | 82.7% (3693/4464) | 74.9% (4266/5694) | 72.1% (3693/5121) | 84.7% (4266/5037) |
| GHH | 0.846 (0.836 to 0.855) | 42.1% (2883/6854) | 79.7% (1743/2188) | 75.6% (3526/4666) | 60.5% (1743/2883) | 88.8% (3526/3971) |
| GAPS (≤21) | 0.737 (0.731 to 0.743) | 45.1% (12220/27078) | 65.8% (6948/10565) | 68.1% (11241/16513) | 56.9% (6948/12220) | 75.7% (11241/14858) |
| BHH | 0.749 (0.739 to 0.758) | 47.6% (4788/10066) | 70.0% (2739/3913) | 66.7% (4104/6153) | 57.2% (2739/4788) | 77.8% (4104/5278) |
| QEHB | 0.755 (0.746 to 0.765) | 47.2% (4799/10158) | 67.5% (3015/4464) | 68.7% (3910/5694) | 62.8% (3015/4799) | 73.0% (3910/5359) |
| GHH | 0.672 (0.659 to 0.685) | 38.4% (2633/6854) | 54.6% (1194/2188) | 69.2% (3227/4666) | 45.3% (1194/2633) | 76.5% (3227/4221) |
| Amb Score (≥6.5) | 0.734 (0.728 to 0.740) | 60.2% (16294/27078) | 83.3% (8802/10565) | 54.6% (9021/16513) | 54.0% (8802/16294) | 83.7% (9021/10784) |
| BHH | 0.724 (0.715 to 0.734) | 62.0% (6242/10066) | 83.8% (3278/3913) | 51.8% (3189/6153) | 52.5% (3278/6242) | 83.4% (3189/3824) |
| QEHB | 0.736 (0.727 to 0.746) | 63.5% (6450/10158) | 85.6% (3820/4464) | 53.8% (3064/5694) | 59.2% (3820/6450) | 82.6% (3064/3708) |
| GHH | 0.735 (0.723 to 0.747) | 52.6% (3602/6854) | 77.9% (1704/2188) | 59.3% (2768/4666) | 47.3% (1704/3602) | 85.1% (2768/3252) |

**Legend.** Results are based on the N=27,078 (N=10,565 with LOS<12) attendances in the validation set for which the three scores were calculable. AUROCs were calculated treating the scores as continuous variables, and predictive accuracy statistics were calculated using the threshold found to maximise the Youden' J statistic when applied to the derivation set. <sup>a</sup> The proportion of attendances that the predictive score would deem suitable for SDEC (i.e. predicted to have LOS<12), using the stated threshold value.

### Evaluation Workshop

The subsequent workshop was attended by 27 people in total, with professional backgrounds including doctors and nurses, those in non-clinical roles, and attendees from England and Scotland, as well as members of the public. The Amb score, GAPS and SDEC-T were discussed, reviewing performance and potential deployment. There was agreement that the inclusion of presenting symptoms was advantageous, as these were felt to influence likely referral to SDEC. The inclusion of a staggered score by age in decades was seen as advantageous rather than the simple cut off used in Amb. Removing the ability to get home independently as a component of the score was also seen as advantageous, as all centres could offer transport if needed. Overall, the workshop agreed that SDEC-T could be deployed in different settings and that performance was appropriate for use.

## Discussion

This paper presents the co-development and validation of a novel risk scoring system, the SDEC-T, designed to predict the likelihood of suitability for treatment in the SDEC setting (using LOS<12 as a surrogate outcome) informed by clinical practice and with input from healthcare professionals, policy makers and patient groups.

As SDEC services have only become as a routine recommended practice within the last ten years, developing from Ambulatory Emergency Care services,(23) and have only recently become a key aspect of strategy within Urgent and Emergency Care (UEC),(6,24) studies and tools available to help select patients for these novel services are limited. A recent scoping review in March 2024 that examined the evidence base for adult medical SDEC in UK NHS Hospitals only identified 18 relevant studies, which were generally of low quality.(25) Reports by the Society for Acute Medicine have described significant variation in the size and staffing models of SDEC units nationally, as well as significant variation in the approaches used to select patients for these services (as identified in the workshops conducted as part of the current study).(26) Unsurprisingly, there is also significant variability in the performance of SDEC nationally, including both the proportion of medical patients who are triaged to these services and those discharged from SDEC.(4)

To our knowledge, this is the first study to propose a predictive score based on a regionally diverse population aimed at supporting an approach to ensure patients are seen in the right place and the right time and specifically for use in SDEC services. A revised assessment of the predictive accuracy of the GAPS and Amb scores to identify patients suitable for SDEC referral (defined as LOS<12 hours) was conducted to establish the current predictive accuracy of these scores. Our centre previously published a similar study based on a single site (QEHB) between 2019-2020, but applied different exclusion criteria.(19) Of these, the major difference was the fact that the previous study excluded attendances with a LOS of more than 48 hours. The rationale for this exclusion was that most of these patients would have been identifiable at triage without the need for a predictive score; hence, such scores would not be applied to this subgroup in practice. The present study considers all ED attendances requiring assessment by the internal medical team. The previous study found predictive accuracy of the GAPS and Amb scores to be poor, with AUROCs of 0.612 and 0.606, compared to 0.741 and 0.733 in the present study.

The reassessment of existing scoring systems revealed that the GAPS score demonstrated marginally superior predictive accuracy compared to the Amb score, with an AUROC of 0.741 (95% CI: 0.738 to 0.744). However, performance varied significantly across different hospital sites. The lowest performance for GAPS was observed at GHH (AUROC of 0.680), while the highest was at QEHB (AUROC of 0.759). This variation may be attributable to the age or socioeconomic differences between the patient populations served by these hospitals; QEHB is located in a more affluent area with lower levels of deprivation compared to GHH. In contrast, the Amb score showed more consistent performance across different sites, with an AUROC range of 0.718 to 0.741.

The newly developed SDEC-T has enhanced the accuracy of predicting short hospital stays and identifying patients suitable for SDEC. A threshold score of ≥9 was determined as the optimal predictor of a LOS<12. SDEC-T demonstrated consistent performance across various hospital sites, with an AUROC range of 0.845–0.858. SDEC-T is highly usable in practice, as it is based on a simple lookup table, which could either be completed manually by clinicians or incorporated into an electronic system. The components of the score are variables that would routinely be recorded at the time of triage in ED; hence, data unavailability should be negligible if the score were calculated prospectively in practice.

However, three components of the score, namely the anticipation of IV treatment, triage category and primary presenting complaint are subject to a degree of subjectivity; hence, inconsistent or unreliable assessments of these factors may impact the performance of the score. Future research should explore the applicability and generalisability of the tool in hospitals beyond the West Midlands region, to assess its broader utility in diverse healthcare settings. In addition, studies using more advanced machine learning/artificial intelligence methodologies to produce more complex tools may be warranted, to assess whether such tools could yield further improved predictive accuracy.

A notable consideration for future research is the operational capacity of SDEC units to handle the potential increase in patient volume indicated by the study’s findings. A total of 45.0% of attendances requiring medical team assessment achieved the surrogate outcome of LOS<12. The use of the suggested threshold for SDEC-T would result in 49.9% of attendances being deemed potentially suitable for SDEC. The practical implications of accommodating such a volume need further investigation, to ensure the operational viability of SDEC services.

### Strengths and Limitations

This study has key strengths, including the large sample size, which allowed for multivariable models to be produced whilst reducing the risk of overfitting. In addition, the cohort comprised data from three hospitals, serving populations with varied demographics, which should improve the generalisability of the findings. The primary limitations of the study related to missing data, which resulted in 11.3% of the cohort being excluded from the analysis of the GAPS and Amb scores, due to the observations required to calculate the MEWS, NEWS and NEWS2 scores being unavailable. This introduced selection bias into the analysis of the scores. Patients where the score were calculable tended to be higher risk, (e.g. older, and more likely to arrive in an ambulance and to be triaged as requiring urgent/immediate care), and so less likely to have LOS<12 than the population as a whole.

However, SDEC-T should not replace the ability of ED staff to triage, treat and send home patients without review by acute and general internal medicine teams, and the patients where NEWS2 was unavailable included a high proportion of patients discharged directly by ED teams. Data were also unavailable for some of the components of the GAPS and Amb scores, meaning that assumptions needed to be made when assigning points. For example, assigning points based on whether patients were prescribed an IV drug within six hours of arrival, rather than whether an IV prescription was anticipated, assumes that the triaging clinician could anticipate the need for IV drugs with 100% accuracy. Since this is unlikely to be the case, the predictive accuracy of this component of the score is likely overestimated in the analysis.

The second limitation related to the outcome used in the analysis. Since it was not possible to retrospectively identify which ED attendances were suitable for SDEC, LOS<12 was used as a surrogate outcome. However, this will have been imperfect, as some patients with longer hospital stays may have been suitable for SDEC, but been subject to delays extending their stay, whilst others may have had shorter stays, but would be better served by being treated in the ED. The final limitation related to the difficulty in retrospectively identifying the cohort eligible for inclusion in the study. In clinical practice, the predictive scores would not be used in patients presenting with complaints that were clearly not suitable for treatment in the SDEC, for example those with fractures or suspected strokes. As such, a range of exclusion criteria were used, which used details of the final diagnoses in the ED, as well as any wards that patients were admitted to during their hospital stay. However, these only gave a broad overview of the reason for each attendance, and so may have incorrectly included or excluded attendances in some cases.

## Conclusion

In conclusion, within this cohort, which is representative of the UK population, SDEC-T has demonstrated greater accuracy in selecting appropriate patients for treatment within SDEC. This enhanced accuracy signifies a significant advancement in the capacity of healthcare trusts to streamline services and ensure rapid, effective patient assessment. Consequently, SDEC-T holds substantial promise for improving patient outcomes and optimising the allocation of healthcare resources in emergency settings.

## Data Availability

To facilitate knowledge in this area, the anonymised participant data, analytical code and a data dictionary defining each field will be available to others through application to PIONEER Data Hub via the corresponding author.

## Study Registration and Protocol

This study was conducted in alignment with the TRIPOD-AI guidelines to ensure transparency in reporting the development and validation of a predictive model for patient suitability in Same Day Emergency Care (SDEC). Although no formal protocol was published for this study, it was registered with PIONEER, the Health Data Research Hub for Acute Care, under the identifier DRF PDR090. This registration outlines the scope, methodology, and ethical considerations of the study, ensuring adherence to best practices in data use and model transparency.

## Author contribution

D. Atkin, S. Gallier, and E. Sapey designed the study, collated data, performed some analysis, and wrote the manuscript. F. Evison curated data and supported statistical analysis. J. Hodson performed and supported statistical analyses. C. Atkin and S. Gallier performed statistical analysis and wrote the first draft of the manuscript. All authors amended the manuscript and approved the final version.

## Acknowledgements

This work was supported by PIONEER, the Health Data Research Hub in acute care, NIHR Midlands Patient Safety Research Collaboration (PSRC) and the NIHR Applied Research Collaboration (ARC) West Midlands and NIHR Birmingham Biomedical Research Centre. Work was also supported by Health Data Research UK. This work uses data provided by patients and collected by the NHS as part of their care and support. We would like to acknowledge the contribution of all staff, key workers, patients and the community who have supported our hospitals and the wider NHS at this time.

## Supplementary Figure Legends

**Supplementary Figure 1.**
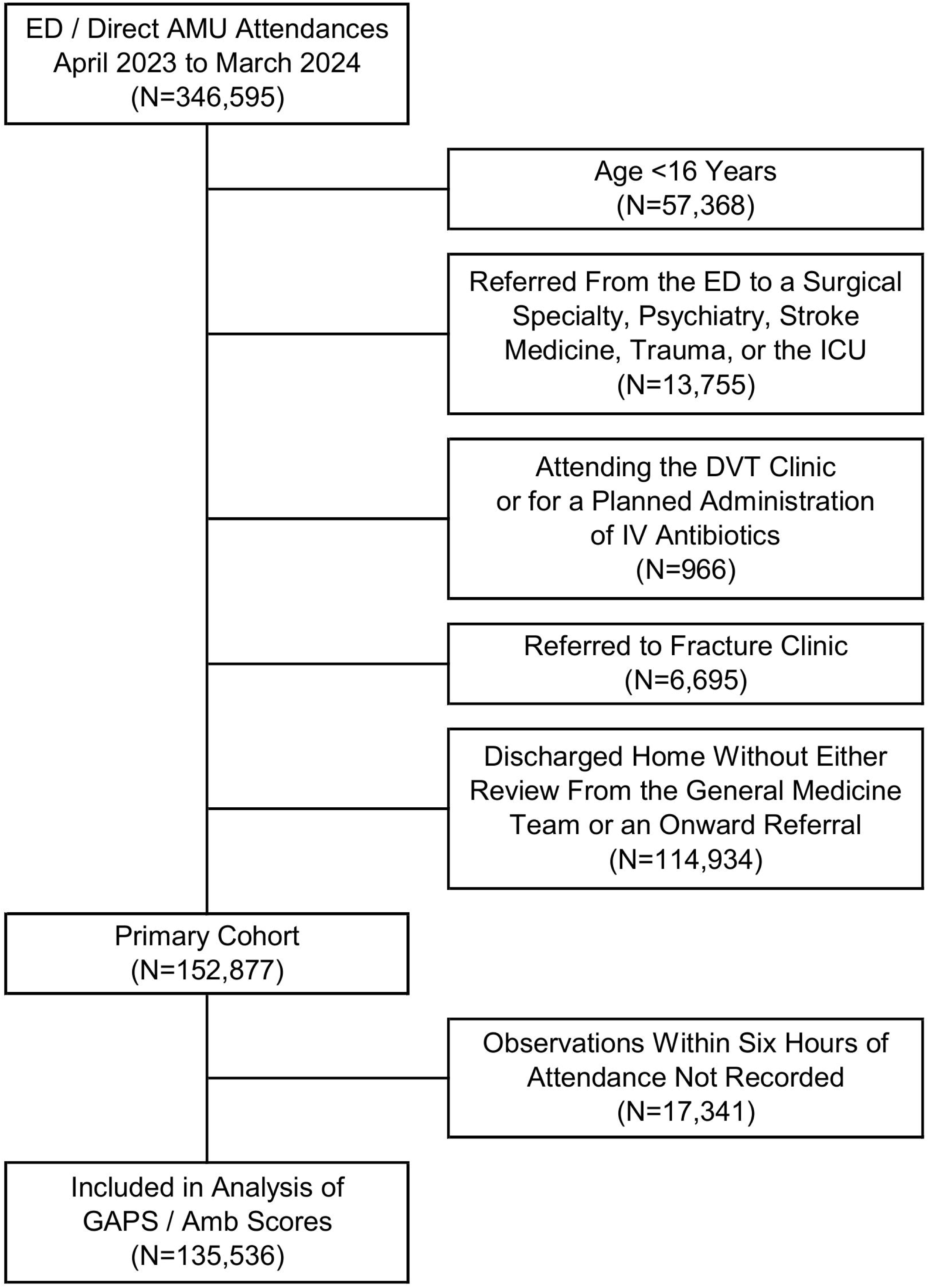
Study flowchart. **Legend**. Amb Score: Ambulatory Score, AMU: Acute Medical Unit, DVT: Deep Vein Thrombosis, ED: Emergency Department, GAPS: Glasgow Admission Prediction Score, ICU: Intensive Care Unit, IV: Intravenous.

**Supplementary Figure 2.**
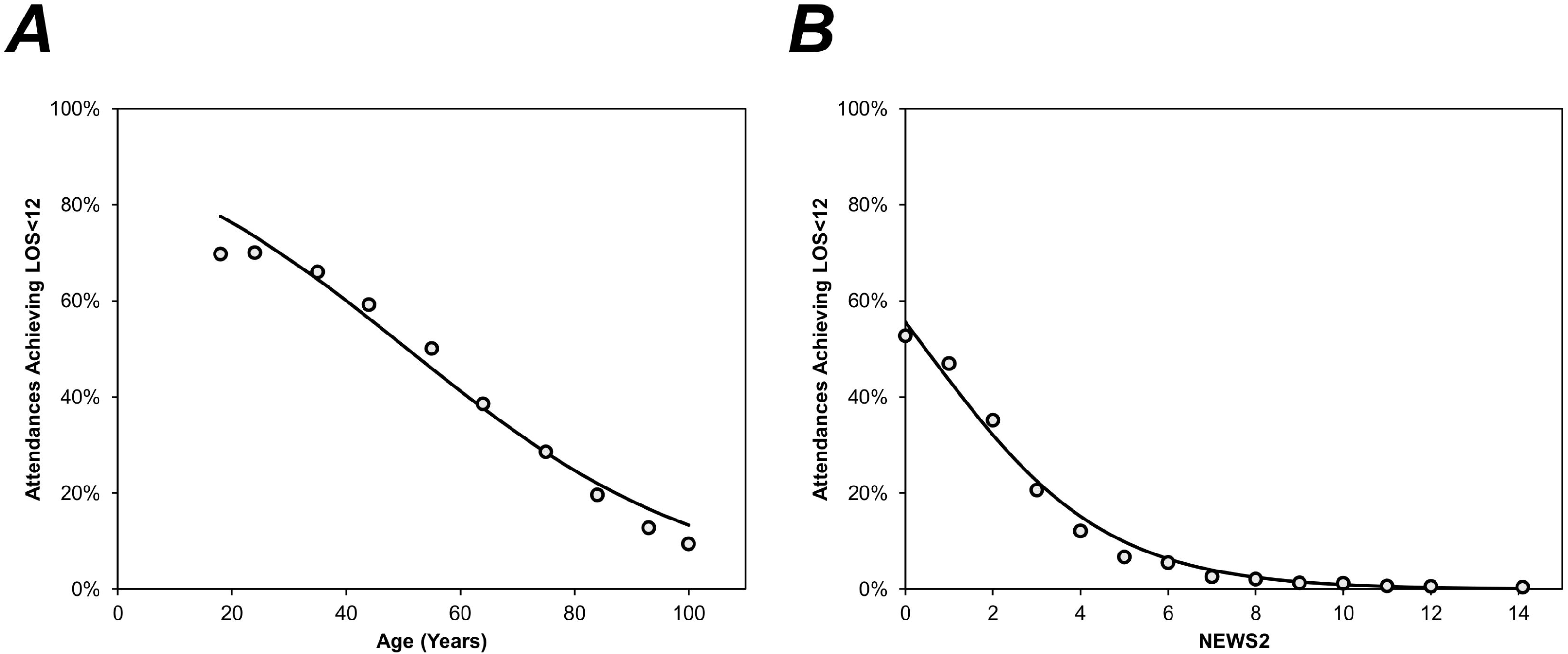
Associations between other risk factors and LOS<12. **Legend**. Plots are based on the attendances in the derivation set. The analysis of age was based on N=122,302; points represent the rates of LOS<12 within each decade of age, and are plotted at the mean age of the interval. The analysis of NEWS2 was based on N=108,451, after excluding cases where this was incalculable; points represent the rates of LOS<12 for each value of the score, with the final point combining scores of 13-20, due to small samples sizes, and plotted at the mean score within this interval. Trend lines are from binary logistic regression models on the admission-level data, with either the age (to the nearest year) or the NEWS2 score as a covariate. LOS<12: Discharged alive with a length of stay of <12 hours. LOS: Length of stay, NEWS2: National Early Warning Score 2

**Supplementary Table 1.**
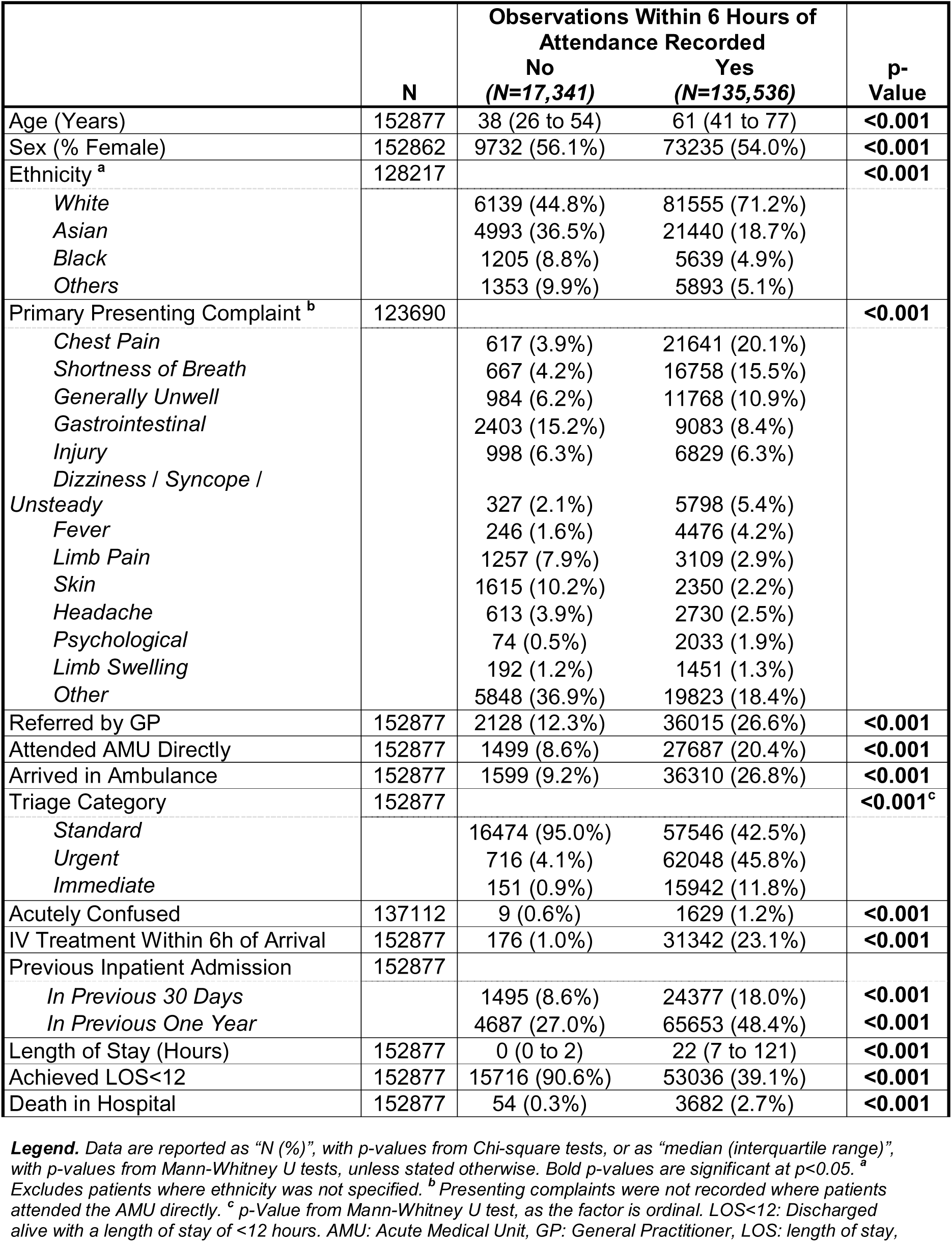
Cohort characteristics in attendances with no observations recorded.

|  |  | Observations Within 6 Hours of Attendance Recorded |  |  |
| --- | --- | --- | --- | --- |
|  | N | No<br>(N=17,341) | Yes<br>(N=135,536) | p-Value |
| Age (Years) | 152877 | 38 (26 to 54) | 61 (41 to 77) | **<0.001** |
| Sex (% Female) | 152862 | 9732 (56.1%) | 73235 (54.0%) | **<0.001** |
| Ethnicity <sup>a</sup> | 128217 |  |  | **<0.001** |
| White |  | 6139 (44.8%) | 81555 (71.2%) |  |
| Asian |  | 4993 (36.5%) | 21440 (18.7%) |  |
| Black |  | 1205 (8.8%) | 5639 (4.9%) |  |
| Others |  | 1353 (9.9%) | 5893 (5.1%) |  |
| Primary Presenting Complaint <sup>b</sup> | 123690 |  |  | **<0.001** |
| Chest Pain |  | 617 (3.9%) | 21641 (20.1%) |  |
| Shortness of Breath |  | 667 (4.2%) | 16758 (15.5%) |  |
| Generally Unwell |  | 984 (6.2%) | 11768 (10.9%) |  |
| Gastrointestinal |  | 2403 (15.2%) | 9083 (8.4%) |  |
| Injury |  | 998 (6.3%) | 6829 (6.3%) |  |
| Dizziness / Syncope / Unsteady |  | 327 (2.1%) | 5798 (5.4%) |  |
| Fever |  | 246 (1.6%) | 4476 (4.2%) |  |
| Limb Pain |  | 1257 (7.9%) | 3109 (2.9%) |  |
| Skin |  | 1615 (10.2%) | 2350 (2.2%) |  |
| Headache |  | 613 (3.9%) | 2730 (2.5%) |  |
| Psychological |  | 74 (0.5%) | 2033 (1.9%) |  |
| Limb Swelling |  | 192 (1.2%) | 1451 (1.3%) |  |
| Other |  | 5848 (36.9%) | 19823 (18.4%) |  |
| Referred by GP | 152877 | 2128 (12.3%) | 36015 (26.6%) | **<0.001** |
| Attended AMU Directly | 152877 | 1499 (8.6%) | 27687 (20.4%) | **<0.001** |
| Arrived in Ambulance | 152877 | 1599 (9.2%) | 36310 (26.8%) | **<0.001** |
| Triage Category | 152877 |  |  | **<0.001<sup>c</sup>** |
| Standard |  | 16474 (95.0%) | 57546 (42.5%) |  |
| Urgent |  | 716 (4.1%) | 62048 (45.8%) |  |
| Immediate |  | 151 (0.9%) | 15942 (11.8%) |  |
| Acutely Confused | 137112 | 9 (0.6%) | 1629 (1.2%) | **<0.001** |
| IV Treatment Within 6h of Arrival | 152877 | 176 (1.0%) | 31342 (23.1%) | **<0.001** |
| Previous Inpatient Admission | 152877 |  |  |  |
| In Previous 30 Days |  | 1495 (8.6%) | 24377 (18.0%) | **<0.001** |
| In Previous One Year |  | 4687 (27.0%) | 65653 (48.4%) | **<0.001** |
| Length of Stay (Hours) | 152877 | 0 (0 to 2) | 22 (7 to 121) | **<0.001** |
| Achieved LOS<12 | 152877 | 15716 (90.6%) | 53036 (39.1%) | **<0.001** |
| Death in Hospital | 152877 | 54 (0.3%) | 3682 (2.7%) | **<0.001** |

**Legend.** Data are reported as “N (%)”, with p-values from Chi-square tests, or as “median (interquartile range)”, with p-values from Mann-Whitney U tests, unless stated otherwise. Bold p-values are significant at $p < 0.05$ . <sup>a</sup> Excludes patients where ethnicity was not specified. <sup>b</sup> Presenting complaints were not recorded where patients attended the AMU directly. <sup>c</sup> p-Value from Mann-Whitney U test, as the factor is ordinal. LOS<12: Discharged alive with a length of stay of <12 hours. AMU: Acute Medical Unit, GP: General Practitioner, LOS: length of stay,

